# Characterising the 2023 chikungunya outbreak in Paraguay and the potential impact of a vaccine campaign

**DOI:** 10.1101/2024.12.02.24318291

**Authors:** Pastor E Pérez-Estigarribia, Gabriel Ribeiro dos Santos, Simon Cauchemez, Cynthia Vazquez, Ana Karina Ibarrola-Vannucci, Guillermo Sequera, Shirley Villalba, María José Ortega, Jose Luis Di Fabio, Danny Scarponi, Christinah Mukandavire, Arminder Deol, Águeda Cabello, Elsi Vargas, Cyntia Fernández, Liz León, Henrik Salje

## Abstract

There now exists a licensed chikungunya vaccine, however, it remains unclear if it could be deployed during outbreaks to reduce the health burden. We used an epidemic in Paraguay as a case study. We conducted a seroprevalence study and used models to reconstruct epidemic transmission dynamics, providing a framework to assess the theoretical impact of a vaccine had it been available. We estimated 33.0% (95% CI: 30.1-36.0) of the population became infected during the outbreak, 6.3% (95%CI: 5.8-6.9) of which were detected by the surveillance system, with a mean infection fatality ratio of 0.013 % (95%CI: 0.012-0.014). A disease-blocking vaccine with 75% efficacy deployed in 40% of > 12-year-olds over a three-month period would have prevented 34,200 (95% CI: 30,900-38,000) cases, representing 23% of cases, and 73 (95%CI: 66-81) deaths. If the vaccine also leads to infection blocking, 88% of cases would have been averted. These findings suggest the vaccine is an important new tool to control outbreaks.

## Introduction

Chikungunya virus (CHIKV) is an alphavirus transmitted by *Aedes* mosquitoes^1^. Outbreaks occur through global tropical and subtropical countries^2^. Infection in humans can cause a range of acute symptoms, including fever, headache and rash. In addition, there are long-term effects of severe arthralgia, which can take months to resolve^3^. Infection can also be deadly, with an estimated 0.1% of cases detected by surveillance systems resulting in death, with elderly age, female sex and comorbidities linked to increased risk of death^4,5^.

Following significant investment, the first chikungunya vaccine, IXCHIQ, was licensed by the FDA at the end of 2023 through an accelerated pathway^6^. In anticipation of the vaccine, Gavi, which funds vaccines in lower-income countries, placed chikungunya vaccines on a Learning Agenda^7^. This means there is currently insufficient evidence to fund the use of the vaccine. Instead, it was felt there needs to be a stronger evidence-base to guide how to use the vaccine. As chikungunya outbreaks are often unpredictable in timing and duration, there remains interest in the development of vaccine stockpiles and only vaccinating populations once an outbreak is detected. Such an approach is used for cholera vaccines^8^. However, it remains unclear if the dynamics of chikungunya epidemics would allow for such a deployment strategy. In particular, responding to an outbreak relies on early detection of the transmission through the surveillance system and then subsequent rollout of the vaccine.

Our understanding of chikungunya epidemiology relies largely on case data reported through surveillance data. However, as clinical misdiagnosis is common, many infected individuals are asymptomatic, or only develop mild symptoms. Further, sick individuals may not visit formal healthcare providers, thus it is difficult to understand the relationship between the reported number of cases and underlying patterns of transmission^9,10^. In this context, seroprevalence studies can be used to reconstruct the underlying infection patterns, and quantify the probability that infected individuals are detected by the surveillance system ^2,11^.

Here, we focus on a large nationwide CHIKV East/Central/South African (ECSA) lineage outbreak in Paraguay that occurred in 2022-23, where there were 142,412 reported cases and 298 deaths^12,13^. While there had been some cases reported in Paraguay each year prior to the outbreak, the largest number of annual cases previously had been 4,509 in 2015, with most years having fewer than 1,000 cases^14^. As we have detailed understanding of the epidemiology, coupled with a new understanding of the underlying infection levels from a seroprevalence study, this outbreak provides an excellent opportunity to understand the impact of a vaccine had it been available at the time.

## Results

From Sep 2022 to Sep 2023, there were 142,412 detected chikungunya cases detected through the national surveillance system, with an average incidence of 208 cases per 10,000 people (2% clinical attack rate). The reported case incidence was greatest in the capital region (Metropolitana, which contains Asuncion) and lowest in Centro Sur (Figure 1A). The incidence of detected cases differed strongly by age and sex, with a steady increase in incidence by age (Figure 1B). Case incidence was 1.43 times higher in females than in males. There was also a strong pattern in deaths by age and sex (Figure 1C), with fatal cases concentrated in the youngest (16.8% of deaths in infants <1y) and the oldest (53.0% of deaths in those >70y), 122 deaths were in females, and 176 deaths were in males.

**Figure 1:**
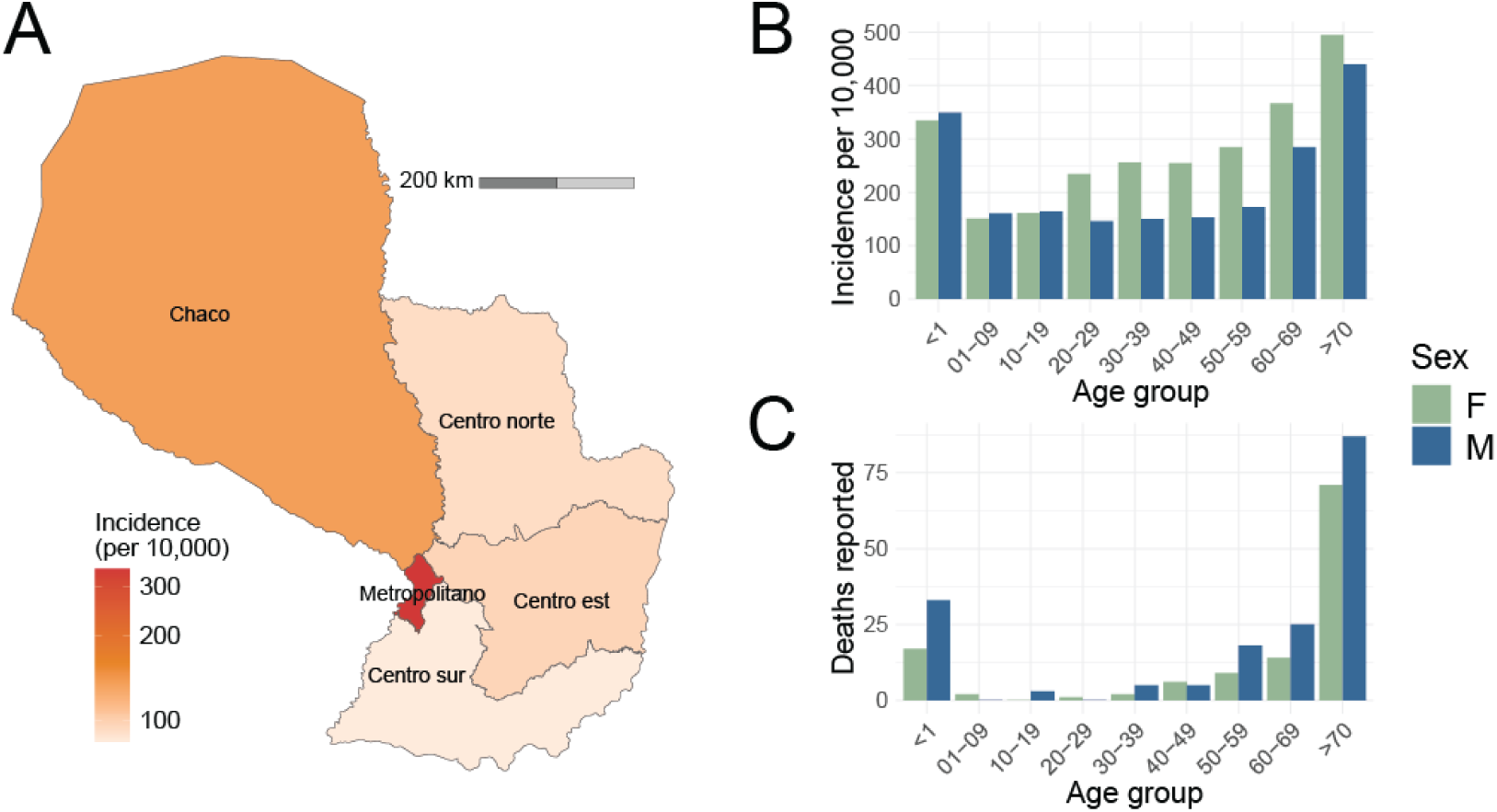
Reported case distribution. **(A)** Map with mean incidence within the five subregions. **(B)** Incidence per age and sex of reported cases. **(C)** Number of deaths by age and sex.

After the end of the outbreak, we tested 1001 individuals who provided sera to blood banks around the country for IgG antibodies to CHIKV (Figure 2A). We found that 340/1001 (34.0% 95%CI: 31.1-37.0) were seropositive. Seropositivity was greatest in the Centro Est region (47.2%, 95%CI: 41.20-53.4) and lowest in the Centro Sur region (7.4% 95%CI: 4.8-11.3) (Figure 2B). Seropositivity did not differ significantly by age (p-value of 0.35) or sex (p-value of 0.08). After accounting for immunity prior to the outbreak, measured at 5% in the capital region from a prior seroprevalence study in 2017 (50 IgG positive individuals from 1,000 tested) and assumed 0% elsewhere, and the distribution of the population across the country, we estimated a mean infection national attack rate of 33.0% (95% CI: 30.1-36.0), equivalent to 2.3 million infections (95%CI: 2.1-2.5 million). We estimated that the surveillance system detected 6.5% (95%CI: 5.9-7.1) of these infections, ranging from 11.3% (95%CI: 7.40-17.6) in Centro Sur to 2.0% (95%CI: 1.8-2.4) in the Centro Est region. The probability of an infected person being detected depended strongly on age and sex, ranging from 4.7% (95%CI: 4.3-5.2) of infected individuals that were 1-9 years old being detected to 14.3% (95%CI: 13.1-15.6) of over >70-year-olds getting detected (Figure 2E, Table S1). On average, females were 1.32 more likely to be detected than males (95%CI; 1.07-1.57). We estimated a mean case fatality ratio of 0.21%, ranging from 0.004% in 20-29 to 1.23% in those >70 years in age. The very youngest were also at high risk of death, with those <1 having a mean IFR of 0.11% (95%CI: 0.10-0.12). The average case fatality ratio was 0.29% in males and 0.15% in females (p-value for difference of <0.001). The mean infection fatality ratio was 0.013% (95%CI: 0.012-0.014%), ranging from 0.00025% (95%CI:0.00023-0.00028) in those 20-29 to 0.18% (95%CI: 0.16-0.29) in those >70-year-olds. We found that the IFR estimates when we assumed the underlying infection attack rate differed by age/sex group as per the seroprevalence study were very similar to when we assumed equal exposure risk across groups (Figure S4).

**Figure 2:**
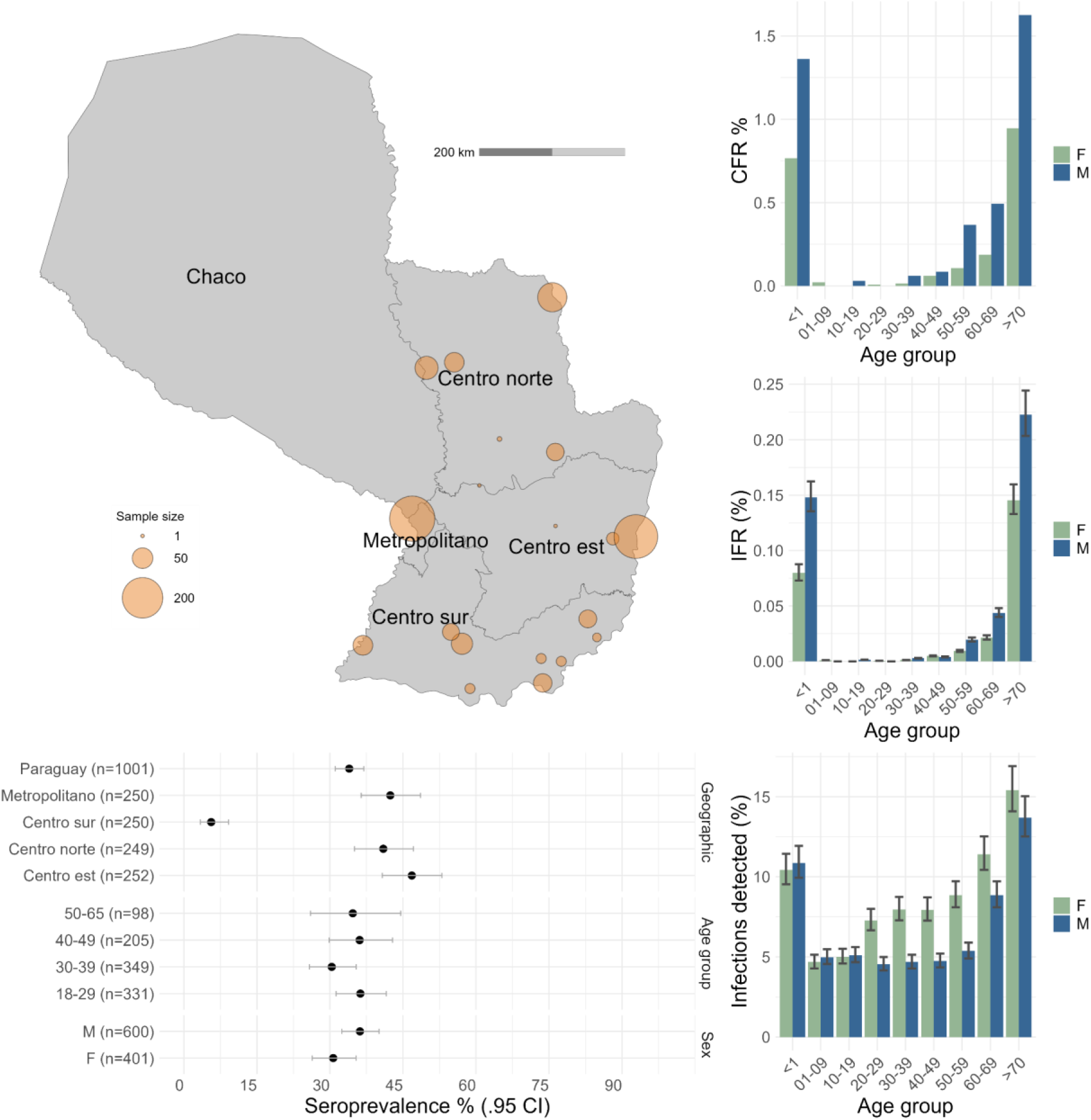
Seroprevalence study and underlying risk of severe disease. **(A)** Location of samples by subregion. **(B)** Seroprevalence by location, age and sex with 95% confidence intervals. **(C)** Case fatality ratio by age and sex. **(D)** Infection fatality ratio by age and sex. **(E)** Probability of severe disease (i.e., being detected by the surveillance system) by age and sex.

To investigate the impact of unreported transmission between 2017 and 2022 leading to greater immunity at the start of the outbreak, we ran a sensitivity analysis assuming 10% of individuals were seropositives in the Metropolitan subregion and 5% in the rest of the country. This led to a slight reduction in the mean infection national attack rate of 27.9% (95%CI: 25.1-30.9), a total number of infections of 1.92 million (95%CI: 1.72-2.12 million) and a mean IFR of 0.016% (95%CI: 0.014-0.017) (Table S2)

We next built a transmission model that jointly estimated the changing transmission patterns over the outbreak and subsequent population immunity. Our model recovered the observed patterns of cases over time and the measured seropositivity (Figure 3A-B). We found that immunity increased sharply to 28.6% (95%CI: 26.5% - 30.9%) nationally 5 months into the epidemic and reached 34.8% (95%CI: 32.0% - 37.7%) after 9 months (Figure 3C). We found that the mean reproductive number over the three first months of the outbreak was 1.9 (95%CI: 1.3 - 2.5) during the first weeks of the outbreak and remained above 1 for 22 weeks (Figure 3D). Overall, 75% of infections occurred over a 29-week period, and 95% of infections occurred over a 35-week period. We found that the effective weekly reproductive number correlated with the country’s mean temperature in that week (Pearson correlation coefficient of 0.75).

**Figure 3:**
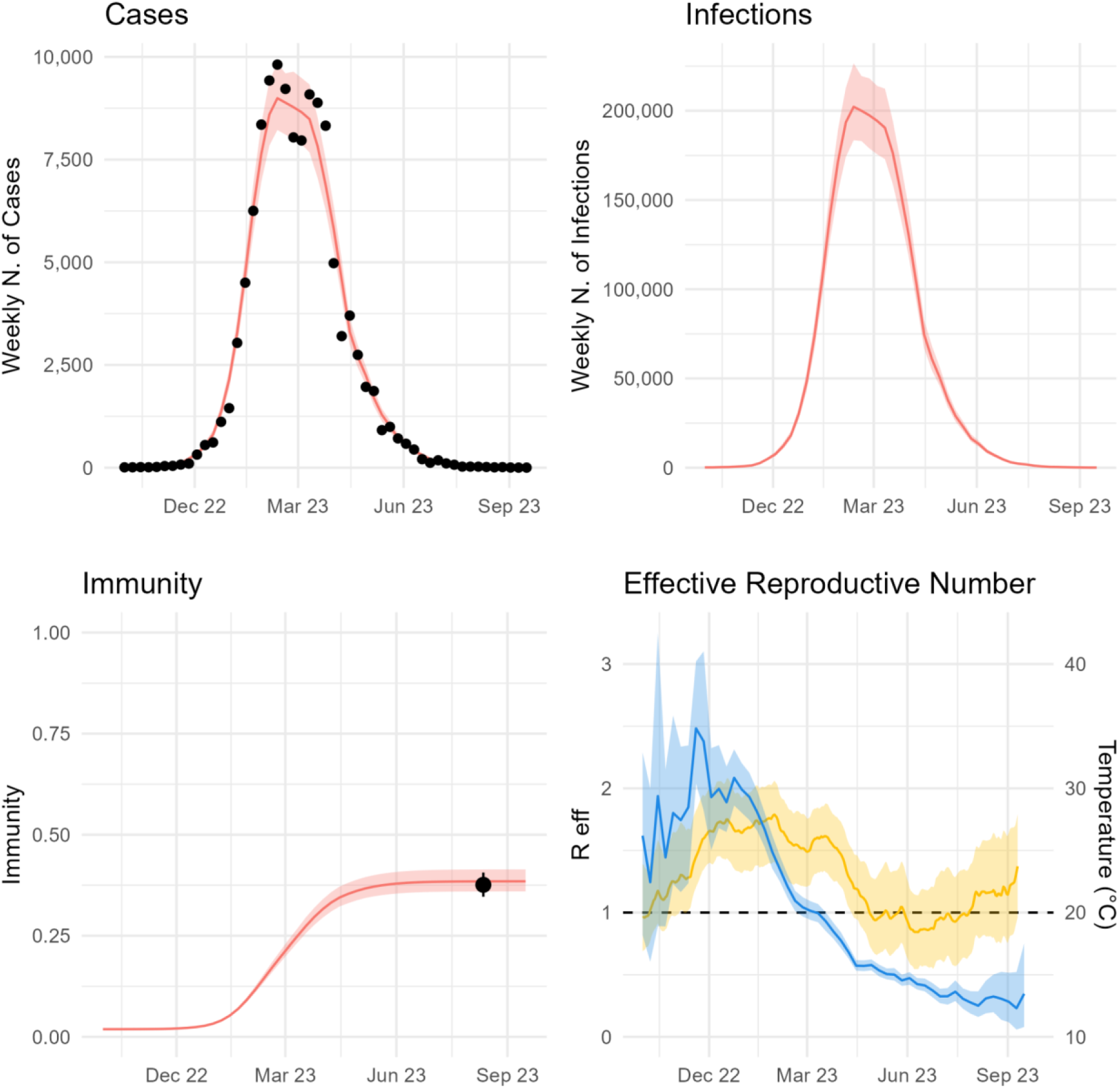
Mathematical model of outbreak. **(A)** Weekly number of cases, black dots is case data from passive surveillance, red is model fit with 95% confidence intervals. **(B)** Weekly number of infections inferred from the model with 95% confidence intervals. **(C)** Evolution of nationwide levels of immunity inferred by the model. Black dot is the result of seroprevalence study. **(D)** Inferred effective reproductive number with 95%CI (blue) and average, minimum and maximum temperature (yellow).

Using our new understanding of the changing underlying transmission dynamics of the virus over the course of the outbreak, we explored the potential impact of a reactive vaccine campaign. To do this, we incorporated an extra compartment into the transmission model for vaccinated individuals, assuming a 75% vaccine efficacy against disease (but did not protect individuals against infection). We then used the fitted transmission parameters to rerun the epidemic but assumed that once the Ministry of Health reported the outbreak (week of 3rd October 2022), a vaccine campaign was initiated in individuals 12 years and over (based on likely initial guidance for the vaccine) and that 40% of the population became vaccinated over a three month period. We used 40% as a benchmark and the rate of rollout based on expert opinion and approximate levels of dengue vaccine (QDenga) uptake during the dengue epidemic in Brazil in 2024. We found that this deployment strategy would have required 2.2 million doses and averted 34,200 (95%CI: 30,500-38,100) cases, including 17,100 (95%CI; 15,500-19,000) cases with chronic sequelae, and 73 (95%CI: 65-81) deaths (Figure 4A, Table S3). This is equivalent to 156 cases, 78 chronic cases and 0.33 deaths averted per 10,000 doses, representing 23% of the cases and deaths that occurred. If, by contrast, only 20% of the population had become vaccinated, then the campaign would have averted 11% of the cases and deaths that occurred (Figure 4B). In a scenario where the vaccine deployment was only initiated 3 months after the outbreak was detected, a campaign with 40% coverage would have averted 13% of the cases and deaths that occurred (Figure 4C). Assuming a higher vaccine efficacy of 98%, results in 31% deaths averted (198 cases and 0.42 deaths averted per 10,000 doses used) (Figure S1).

**Figure 4:**
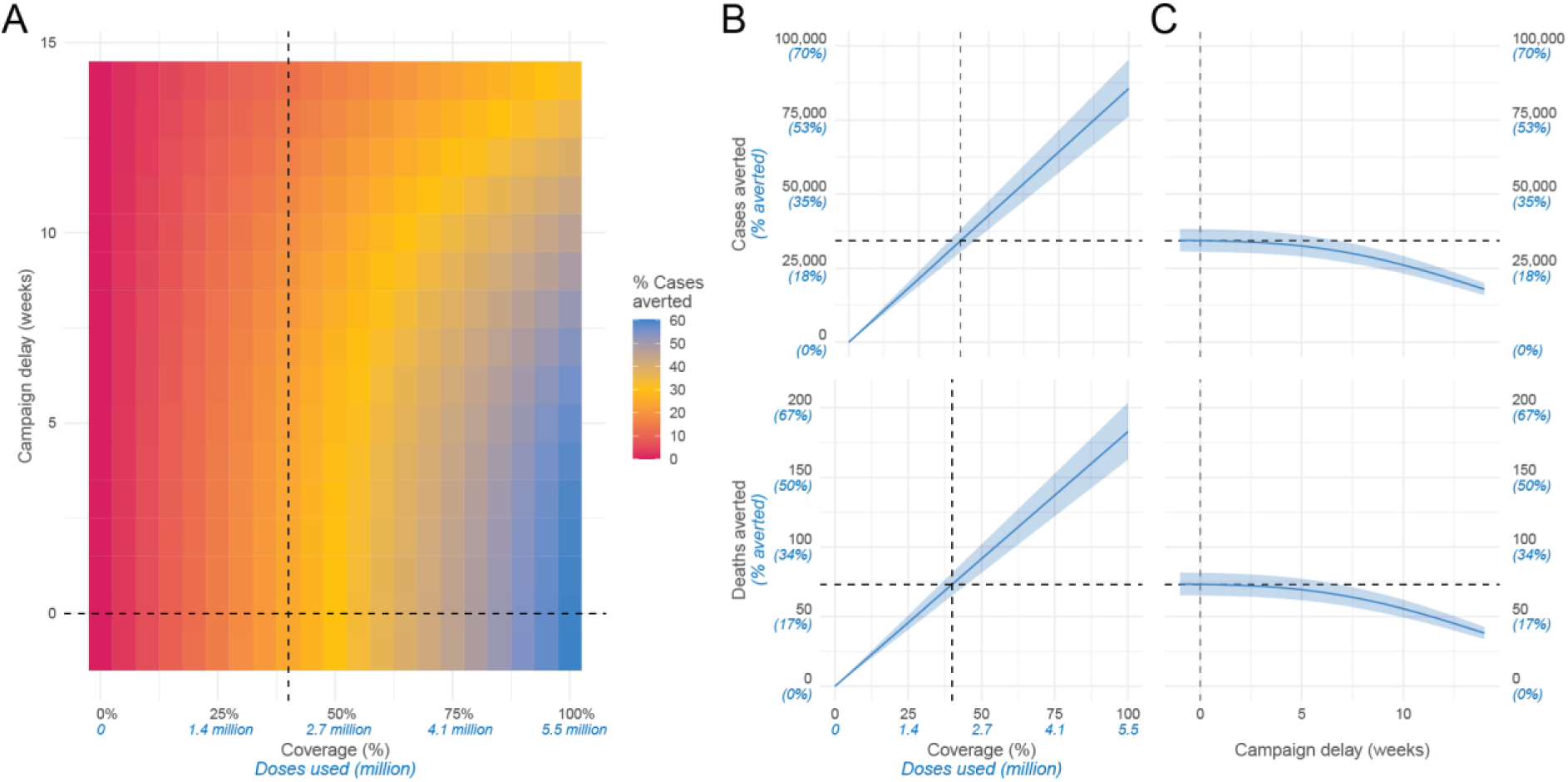
Results of vaccine model. **(A)** Proportion of cases averted for different values of coverage and delay. The dashed black line shows the base case scenario of 40% coverage with no delay between outbreak start and campaign vaccination. **(B)** Infections (top row), cases (middle row) and deaths (bottom row) averted when varying coverage and delay are fixed at 0 weeks. **(C)** Infections (top row), cases (middle row) and deaths (bottom row) averted when varying the delay between the detection of the outbreak and the start of the vaccine campaign, fixing coverage at 40%.

These initial results cautiously assumed that the vaccine prevents disease only, but that vaccinated individuals can still become infected. To assess the impact of the vaccine, if it also blocks onward transmission, consistent with what we believe for other live attenuated vaccines and the observed relationship between titers and risk of infection and disease in chikungunya cohorts, we amended our model to prevent onward spread from infected vaccinated individuals, keeping everything else unchanged (Figure S2) ^15^. We found that a 40% coverage in the same population with 75% efficacy against both infection and disease would have a substantial impact on the epidemic, with 88% deaths averted (573 cases and 1.20 deaths averted per 10,000 doses used).

## Discussion

We have used a large CHIKV outbreak in Paraguay to quantify the underlying detection probabilities of surveillance systems for chikungunya, the attack rate of the epidemic, and to quantify the potential impact of a reactive vaccine campaign. We found that the vaccine could significantly reduce the burden of the virus, highlighting the important potential of this new technology.

Our findings provide an avenue for how to implement this new vaccine. Chikungunya epidemiology remains highly unpredictable in most settings. While some countries have evidence of endemic transmission at the national level, the majority of locations experience infrequent outbreaks, and there may be limited appetite to initiate vaccine campaigns when the near term burden remains highly uncertain and the durability of protection remains unclear^5,16^. By demonstrating that it can be used in a reactive manner, it provides an evidence base for the establishment of stockpiles that can be readily deployed once an outbreak is detected. Overall, the outbreak lasted around 8 months beyond the declaration of the outbreak, with the reduction in the force of infection probably linked to climatic conditions coupled with increased immunity in the population.

This study highlights how sensitive surveillance systems are critical for effective infectious disease response, and should be prioritised for investment. In particular, reactive vaccine campaigns rely on surveillance systems detecting new outbreaks quickly. Due to the similarity in symptoms with dengue and other pathogens and limited access to testing, entire CHIKV epidemics can be missed^11,17^. However, in some countries within South America such as Brazil and Paraguay and potentially elsewhere, which only experienced their first outbreaks in 2014, surveillance systems seem to be better equipped to detect CHIKV outbreaks. The Ministry of Health declared an outbreak in October 2022, when only around 30 cases had been detected. The optimal number of cases threshold that should be used to trigger a vaccine response is unclear and requires careful thought as ultimately there is a trade-off between timely and appropriate response to outbreaks and limiting false alarms. It is likely that the optimal threshold will be setting-specific and will depend on surveillance capabilities and response capabilities.

The accelerated pathway to FDA approval has meant we have a product that is close to being used. However, the absence of traditional phase III trials, does mean we continue to have a limited understanding of the efficacy of the vaccine. However, antibody data from the vaccine are very promising, with high, sustained titers in vaccinated individuals^15,18^. We would also expect live attenuated viruses to act much like a natural infection and, therefore, result in infection blocking. Our sensitivity analysis showing a much larger impact than those based on disease blocking only, may therefore be reasonable.

While the major focus of chikungunya vaccines remains averting morbidity and long-term chronic sequelae resulting from CHIKV infection^2^, there is a growing realisation that chikungunya is a deadly disease ^4^. Nearly 300 individuals lost their lives over the Paraguay outbreak with a mean CFR of 0.21%, higher than previous estimates from the Americas, which range between 0.05% and 0.13% ^19^. The reasons why the CFR was higher are unclear. It seems unlikely that viral factors are linked to the increased fatality as the same ECSA strain has been found elsewhere in the Americas. Instead the higher than expected CFR may be linked to reduced case detection during this large outbreak, with only around one in 20 infections ultimately detected, potentially down to the swamping of the surveillance system. We estimated an IFR of ∼0.01%. Unlike CFRs, IFR estimates are robust to underlying biases in case surveillance. We are not aware of other studies that have directly quantified the underlying CHIKV IFR as the underlying level of infection has rarely been estimated.

Our study has also provided insight into the underlying disease patterns in infected populations. We found a strong effect of age on the probability of detection by the surveillance system. This is consistent with other reports of increasing probability of symptoms by age following infection^5^. We also found a significant probability of detection in females as compared to males. This pattern has been observed elsewhere, including in Brazil and Bangladesh^5,20,21^. Earlier work had suggested higher levels of cases in females may be linked to underlying differences in risk behaviour, with females more likely to be in and around the home, where *Aedes* mosquitoes are typically found. However, we found in this setting there were similar levels of underlying infection between males and females, suggesting that there are increasing probabilities of the development of symptoms in females as compared to males following infection. Differential healthcare-seeking behaviours between males and females may also contribute to this observation. By contrast, we found that the probability of death was significantly higher in males than females. These complex differences in the development of symptoms and risk of severed disease between the sexes is potentially linked to complex immunological factors, consistent with that found across different infectious diseases^22^. Given the strong patterns of symptoms and risk of death by age, the elderly should be prioritised for vaccination.

Our study is subject to limitations. We used blood bank sera in our seroprevalence study, which means we have only adults in this study and little data on participants. Individuals who provide blood may have a different underlying risk of chikungunya than the rest of the population; however, given the high attack rate in the country, any effect is likely to be minimal. Our seroprevalence study may also be affected by false-positives due to cross-reactivity with Mayaro virus. However, we expect it would only have a minimal impact on the estimated overall number of CHIKV infected individuals as the vector that transmits Mayaro is mostly found in sparsely populated forested regions where the *Haemagogus* mosquito resides^21^. The applicability of our results to other regions where the chikungunya virus circulates is unclear, especially as the outbreak was detected relatively quickly in Paraguay. Finally, the outbreak in Paraguay was large, infecting around 3 in 10 individuals. The impact of the vaccine in smaller outbreaks may not be as significant. Nevertheless, the largest outbreaks are ultimately the most disruptive to public health and general healthcare provision. Therefore a significant reduction in the impact of these major epidemics is a major step forward on our battle against the virus. .

With explosive outbreaks, chikungunya virus remains a threat to public health across large areas of the globe. However, we now have a tool to combat these outbreaks and, following this work, an evidence base to support the rapid deployment of the vaccine following outbreak detection. The development of appropriate regional stockpiles, robust vaccine deployment protocols, and local vaccine licensure will be a necessary next step in using the vaccine.

## Methods

### Epidemic case data

We used weekly case data from Sep 2022 to Sep 2023. All suspected chikungunya cases were reported to the Dirección General de Vigilancia de la Salud, Ministerio de Salud Pública y Bienestar Social de Paraguay. Around 60% of suspected cases undergo confirmatory testing through PCR, with an additional 23% tested through ELISA IgM. Suspected infections are defined as any person with sudden onset of fever and sudden onset of arthralgia or disabling arthritis unexplained by another medical condition. Probable infections are defined as any suspected case with a positive laboratory result for CHIKV (IgM ELISA) or any suspected case of CHIKV with an epidemiological link to a confirmed case. Confirmed infections are suspected or probable CHIKV cases involving real-time RT-PCR or viral isolation^12,23^.

### New seroprevalence study

We conducted a seroprevalence study using blood bank serum in four of five subregions (defined by the Ministry of Health, “Ejes”) of Paraguay (Metropolitana, Centro Sur, Centro Este and Centro Norte). We were not able to obtain sera from the final subregion (Chaco). In each subregion, we worked with the local blood collection service. Samples were collected from 25 July 2023 to 23 August 2023 from people aged 18-65 years attending a blood donation service, with a target sample size of 250 people per Axis. We used IgG Euroimmun ELISA kits to test the samples for evidence of IgG antibodies to Chikungunya virus. The testing was conducted in the Laboratorio Central de Salud Pública, Paraguay. We adjusted our seroprevalence estimates to account for the 98.6% sensitivity and 98% specificity of the kits.

### Statistical analyses

#### Age- and sex-specific probability of disease

We assumed that outside the capital subregion (Metropolitana), all individuals were susceptible prior to the outbreak. In the capital subregion, we assumed that 5% of individuals were seropositive prior to the outbreak, based on a household-based seroprevalence study in 1,000 individuals aged 5-65 conducted in 2017 (personal communication, Paraguay Ministry of Public Health, Figure S3). Uncertainty on seroprevalence was accounted for by estimating the 95% confidence interval around point estimates assuming a binomial distribution on samples tested. Seroincidence was calculated as the difference between sensitivity-adjusted seroprevalence post-outbreak and seroprevalence at baseline. Uncertainty from the seroprevalence estimates was propagated to the estimates on the underlying number of infections, on the IFR and on probability of detection.

Given non-significant differences in seroprevalence estimates across sex and age groups, we assumed equal risk of exposure by age and estimated the age- and sex-specific number of infections by subregion using the attack rate and demographic data from the national census. We then estimated the probability of disease by dividing the total number of cases by age and sex strata by the estimated number of infections in the strata. We separately estimated the age- and sex-specific infection fatality ratio by dividing the number of deaths in each age and sex strata by the estimated number of infections in the same strata. (Table S1) Uncertainty from the seroprevalence estimates was propagated to the estimates on the underlying number of infections, on the IFR and on probability of detection.

#### Epidemic model

To characterise the chikungunya epidemic trajectory, we developed a compartmental SIR model in which the transmission rate beta varies over time. Weekly transmission rates were assumed to be independent and estimated as free parameters. We assumed the generation time for CHIKV (defined as the average time interval between consecutive infections) was two weeks^20^.

We modelled the number of incident cases, assuming a negative binomial observation process. The likelihood of observing c_obs(t) incident cases on week t given the expected number of infections i_exp(t) for that week is given by the density of a Negative Binomial distribution:

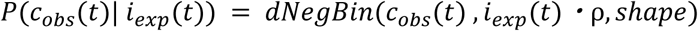

where ⍴ is the detection probability and *shape* is the overdispersion parameter of the negative binomial distribution.

Within the same analytical framework using a joint likelihood, we incorporated the observed number of seropositive individuals in our seroprevalence study, assuming a binomial observation process. The likelihood of observing n_pos(t) positive samples out of n_tot(t) on week t given the expected proportion of susceptible individuals in the population s(t) for that week is given by the density of a Binomial distribution:

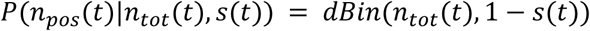

The number of infectious individuals at the start of the outbreak (first week of November 2022) was fixed at 100. The detection probability is the probability that the surveillance system detected an infected individual and was estimated as a free parameter. Parameters were estimated using Markov Chain Monte Carlo (MCMC) with a Metropolis-Hastings algorithm. We used 4 chains of 25,000 iterations, including a burn-in phase of 10,000 iterations and sampling with uniform non-informative priors. Effective sample sizes and R-hat were computed for each parameter. (Table S4)

#### Estimating the impact of the vaccine

To estimate the potential effect of a vaccine, we investigated a scenario where a vaccine was available during the outbreak by adjusting our transmission model. We assumed the vaccine was a leaky vaccine, so that vaccinated individuals had a probability of nevertheless getting infected. The target population was individuals 12 years old and older, consistent with the planned initial target age group of the IXCHIQ vaccine. We initially assumed a vaccine efficacy of 0.75, meaning 75% of vaccinated individuals were protected against disease but not protected against infection. We assumed a delay of two weeks from vaccination to acquisition of immunity^24,25^. We considered a reactive roll-out campaign that started the first week of October 2023. This date represents the moment when the Ministry of Health officially reported the outbreak. We then assumed it takes three months to reach a coverage of 40% of the target population, assuming a fixed weekly rate of vaccination.

We conducted a sensitivity analysis where we varied (a) the level of vaccination coverage and (b) allowed for different delays between the start of the outbreak and the start of the campaign deployment. We also included a sensitivity analysis where we assumed a vaccine efficacy of 95%. Finally, we conducted a separate analysis where we assumed the vaccine blocked individuals from onward transmission. For each scenario, we measured the number of infections, cases and deaths averted as well as the number of doses used.

## Data Availability

All data produce in the present work are contained in the manuscript

https://github.com/Pastor-E-Perez-Estigarribia/ePY

## Supplementary materials

### Supplementary text-SIRV ODE system

The outbreak was characterised using an SIR compartmental model. To estimate the potential impact of a vaccine it was estimated adding vaccinated compartments to the SIR system. We modelled vaccine efficacy (VE) as being a leaky vaccine, with all recipients of the doses receiving partial protection. The set of equations is as follow:

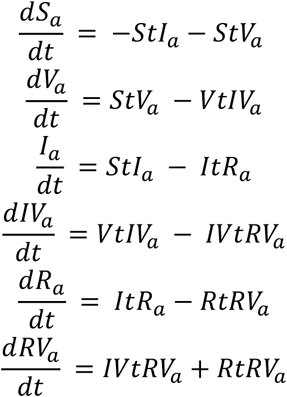

With :

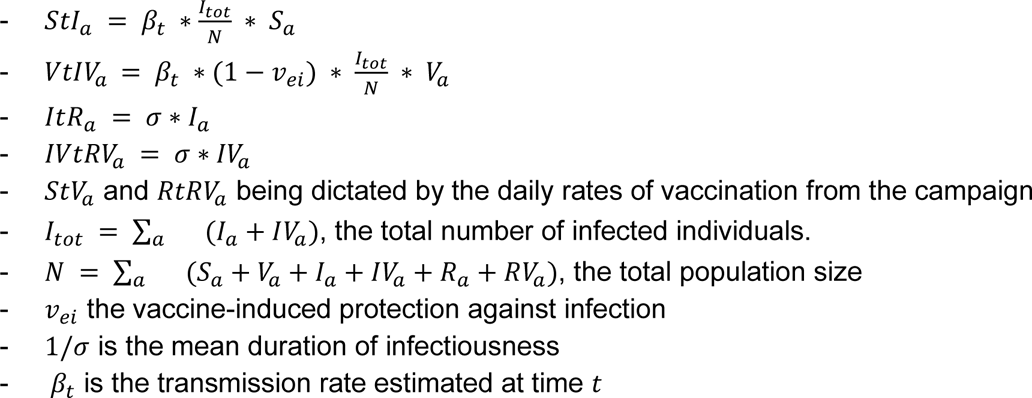

**Table S1:** Statistics stratified by sex, subregion and age groups.

| Group | Cases reported | Deaths reported | Sero-prevalence | Infection attack rate | Nb. estimated infections | IFR (%) | Detection probability | CFR (%) |
| --- | --- | --- | --- | --- | --- | --- | --- | --- |
| **SEX** |  |  |  |  |  |  |  |  |
| **Female** | 81,485 | 122 | 34.8<br>[31.9-37.8] | 33 [30.1-36] | 1,130,000<br>[1,030,000-1,230,000] | 0.0108<br>[0.00992-0.0119] | 0.072<br>[0.066-0.079] | 0.15 |
| **Male** | 60,927 | 176 | 34.8<br>[31.9-37.8] | 33 [30.1-36] | 1,130,000<br>[1,030,000-1,240,000] | 0.0155<br>[0.01420-0.0170] | 0.054<br>[0.050-0.059] | 0.29 |
| **SUBREGION** |  |  |  |  |  |  |  |  |
| **Centro est** | 21,075 | 58 | 47.20<br>[41.20-53.4] | 47.20<br>[41.20-53.4] | 1,040,000 [908,000-1,180,000] | 0.00557<br>[0.00493-0.00639] | 0.020<br>[0.018-0.023] | 0.275 |
| **Centro norte** | 10,399 | 14 | 41.60<br>[35.60-47.8] | 41.60<br>[35.60-47.8] | 476,000 [408,000-547,000] | 0.00294<br>[0.00256-0.00343] | 0.0219<br>[0.0190-0.0255] | 0.135 |
| **Centro sur** | 9,494 | 18 | 7.41 [4.77-11.3] | 7.41 [4.77-11.3] | 83,800 [53,900-128,000] | 0.02150<br>[0.01400-0.03340] | 0.1130<br>[0.0740-0.1760] | 0.19 |
| **Chaco** | 3,05<br>1 | 2 | - | - | - | - | - | 0.06<br>56 |
| **Metropolitano** | 98,6<br>65 | 206 | 43.00<br>[37.00-49.2] | 41.10<br>[35.10-47.3] | 1,170,00<br>0<br>[1,000,0<br>00-1,350,00<br>0] | 0.017<br>60<br>[0.015<br>30-0.020<br>60] | 0.0842<br>[0.0732-0.0986] | 0.20<br>9 |
| **AGE GROUP** |  |  |  |  |  |  |  |  |
| **<1** | 4,64<br>1 | 50 | 34.8<br>[31.9-37.8] | 33 [30.1-36] | 44,800 [40,800-48,800] | 0.112<br>000<br>[0.102<br>000-0.122<br>000] | 0.1040<br>[0.0950-0.1140] | 1.08 |
| **01-09** | 18,8<br>28 | 2 | 34.8<br>[31.9-37.8] | 33 [30.1-36] | 399,000<br>[364,000<br>-<br>436,000] | 0.000<br>501<br>[0.000<br>459-0.000<br>549] | 0.0471<br>[0.0432-0.0517] | 0.01<br>06 |
| **10-19** | 19,8<br>54 | 3 | 34.8<br>[31.9-37.8] | 33 [30.1-36] | 403,000<br>[367,000<br>-<br>439,000] | 0.000<br>745<br>[0.000<br>683-0.000<br>817] | 0.0493<br>[0.0452-0.0541] | 0.01<br>51 |
| **20-29** | 22,6<br>89 | 1 | 34.8<br>[31.9-37.8] | 33 [30.1-36] | 395,000<br>[361,000<br>-<br>431,000] | 0.000<br>253<br>[0.000<br>232-0.000<br>277] | 0.0574<br>[0.0526-0.0629] | 0.00<br>441 |
| **30-39** | 21,9<br>39 | 7 | 34.8<br>[31.9-37.8] | 33 [30.1-36] | 357,000<br>[326,000<br>-<br>390,000] | 0.001<br>960<br>[0.001<br>800-0.002<br>150] | 0.0614<br>[0.0563-0.0673] | 0.03<br>19 |
| **40-49** | 15,6<br>73 | 11 | 34.8<br>[31.9-37.8] | 33 [30.1-36] | 254,000<br>[232,000<br>-<br>277,000] | 0.004<br>330<br>[0.003<br>970-0.004<br>750] | 0.0617<br>[0.0566-0.0677] | 0.07<br>02 |
| **50-59** | 13,4<br>04 | 27 | 34.8<br>[31.9-37.8] | 33 [30.1-36] | 192,000<br>[176,000<br>-<br>210,000] | 0.014<br>000<br>[0.012<br>900-0.015<br>400] | 0.0696<br>[0.0638-0.0763] | 0.20<br>1 |
| **60-69** | 12,5<br>16 | 39 | 34.8<br>[31.9-37.8] | 33 [30.1-36] | 126,000<br>[115,000<br>-<br>137,000] | 0.031<br>000<br>[0.028<br>400-0.034<br>000] | 0.0995<br>[0.0912-0.1090] | 0.31<br>2 |
| **>70** | 12,8<br>68 | 158 | 34.8<br>[31.9-37.8] | 33 [30.1-36] | 90,200 [ 82,300-98,500] | 0.175<br>000<br>[0.160<br>000-0.192<br>000] | 0.1430<br>[0.1310-0.1560] | 1.23 |
| **TOTAL** |  |  |  |  |  |  |  |  |
| **Total** | **142,**<br>**412** | **298** | **34.8**<br>**[31.9-**<br>**37.8]** | **33 [30.1-**<br>**36]** | **2,260,00**<br>**0**<br>**[2,060,0**<br>**00-**<br>**2,470,00**<br>**0]** | **0.013**<br>**2**<br>**[0.01**<br>**21-**<br>**0.014**<br>**4]** | **0.063**<br>**[0.0577-**<br>**0.069]** | **0.20**<br>**9** |

**Table S2.** : Sensitivity analysis comparing scenario assuming 5% initial seroprevalence in the capital and 0% elsewhere compared to 10% in the capital city and 10% elsewhere.

| Scenario | Initial seroprevalence | Infection attack rate | Number of estimated infections | IFR (%) | Detection probability |
| --- | --- | --- | --- | --- | --- |
| Base case | 1.85% | 33 [30.1-36] | 2,260,000<br>[2,060,000-2,470,000] | 0.0132 [0.0121-0.0144] | 0.063 [0.0577-0.069] |
| Higher seroprevalence | 6.87% | 27.9 [25.1-30.9] | 1,920,000<br>[1,720,000-2,120,000] | 0.0155 [0.014-0.0173] | 0.0743 [0.0671-0.0828] |

**Table S3:** Vaccine impact under different scenarios.

| Scenario | Doses used | Infections averted | Cases averted | Cases presenting chronic sequelae averted | Deaths averted |
| --- | --- | --- | --- | --- | --- |
| **Base case** | 2,190,000 | 618,000<br>(95%CI: 558,000-686,000) | 34,200 (95%CI: 30,900- 38,000) | 17,100 (95%CI: 15,500-19,000) | 73 (95%CI: 66- 81) |
| **Coverage 20%** | 1,100,000 | 309,000<br>(95%CI: 279,000-343,000) | 17,100 (95%CI: 15,500- 19,000) | 8,560 (95%CI: 7,730- 9,500) | 37 (95%CI: 33- 41) |
| **Higher baseline seroprevalence** | 2,190,000 | 359,000<br>(95%CI: 245,000-433,000) | 29,600 (95%CI: 20,200- 35,700) | 14,800 (95%CI: 10,100-17,800) | 63 (95%CI: 43- 76) |
| **98% efficacy against disease** | 2,190,000 | 808,000<br>(95%CI: 729,000-896,000) | 44,800 (95%CI: 40,400- 49,600) | 22,400 (95%CI: 20,200-24,800) | 96 (95%CI: 86-106) |
| **Infection blocking** | 2,190,000 | 2,340,000<br>(95%CI: 2,190,000-2,510,000) | 124,000 (95%CI: 116,000-132,000) | 61,800 (95%CI: 58,000-66,200) | 259 (95%CI: 243-277) |

**Table S4:** MCMC diagnostics - Prior distribution, posterior estimates, effective sample size and R-hat for all parameters estimated with the transmission model.

| Parameter | Prior distribution | Posterior median and 95% quantile | Effective Sample Size | R-hat |
| --- | --- | --- | --- | --- |
| **Reporting** | Uniform [0,1] | 0.0444<br>(95%CI:<br>0.0403-0.048) | 605 | 1.12 |
| **Transmission rate - week 01** | Uniform [0,10] | 0.693<br>(95%CI:<br>0.293-1.39) | 33.1 | 1.42 |
| **Transmission rate - week 02** | Uniform [0,10] | 0.47 (95%CI:<br>0.149-1.22) | 51 | 3.59 |
| **Transmission rate - week 03** | Uniform [0,10] | 0.872<br>(95%CI:<br>0.349-2.46) | 28 | 3.28 |
| **Transmission rate - week 04** | Uniform [0,10] | 0.692<br>(95%CI:<br>0.193-1.37) | 31.8 | 1.92 |
| **Transmission rate - week 05** | Uniform [0,10] | 0.581<br>(95%CI:<br>0.186-1.04) | 55.5 | 2.45 |
| **Transmission rate - week 06** | Uniform [0,10] | 0.513<br>(95%CI:<br>0.266-0.875) | 68.3 | 1.19 |
| **Transmission rate - week 07** | Uniform [0,10] | 0.991<br>(95%CI:<br>0.457-1.53) | 18.2 | 2.53 |
| **Transmission rate - week 08** | Uniform [0,10] | 0.73 (95%CI: 0.393-1.19) | 51.4 | 1.91 |
| **Transmission rate - week 09** | Uniform [0,10] | 0.727 (95%CI: 0.469-1.17) | 24 | 1.34 |
| **Transmission rate - week 10** | Uniform [0,10] | 0.882 (95%CI: 0.556-1.35) | 44.7 | 1.95 |
| **Transmission rate - week 11** | Uniform [0,10] | 0.85 (95%CI: 0.545-1.16) | 88.5 | 1.75 |
| **Transmission rate - week 12** | Uniform [0,10] | 1.18 (95%CI: 0.935-1.65) | 69.5 | 1.52 |
| **Transmission rate - week 13** | Uniform [0,10] | 1.12 (95%CI: 0.795-1.42) | 124 | 1.2 |
| **Transmission rate - week 14** | Uniform [0,10] | 0.896 (95%CI: 0.731-1.1) | 220 | 1.15 |
| **Transmission rate - week 15** | Uniform [0,10] | 0.962 (95%CI: 0.819-1.11) | 239 | 1.07 |
| **Transmission rate - week 16** | Uniform [0,10] | 0.898 (95%CI: 0.79-1.03) | 363 | 1.02 |
| **Transmission rate - week 17** | Uniform [0,10] | 0.999<br>(95%CI:<br>0.895-1.11) | 439 | 1.01 |
| **Transmission rate - week 18** | Uniform [0,10] | 0.959<br>(95%CI:<br>0.862-1.06) | 615 | 1.01 |
| **Transmission rate - week 19** | Uniform [0,10] | 0.933<br>(95%CI:<br>0.845-1.02) | 696 | 1.01 |
| **Transmission rate - week 20** | Uniform [0,10] | 0.891<br>(95%CI:<br>0.814-0.974) | 1010 | 1.01 |
| **Transmission rate - week 21** | Uniform [0,10] | 0.84 (95%CI:<br>0.768-0.911) | 1290 | 1.01 |
| **Transmission rate - week 22** | Uniform [0,10] | 0.767<br>(95%CI:<br>0.706-0.837) | 1970 | 1 |
| **Transmission rate - week 23** | Uniform [0,10] | 0.711<br>(95%CI:<br>0.657-0.767) | 1770 | 1 |
| **Transmission rate - week 24** | Uniform [0,10] | 0.656<br>(95%CI:<br>0.608-0.709) | 1800 | 1 |
| **Transmission rate - week 25** | Uniform [0,10] | 0.618<br>(95%CI:<br>0.572-0.666) | 3090 | 1.01 |
| **Transmission rate - week 26** | Uniform [0,10] | 0.61 (95%CI: 0.565-0.658) | 2170 | 1.02 |
| **Transmission rate - week 27** | Uniform [0,10] | 0.609 (95%CI: 0.564-0.657) | 5320 | 1.02 |
| **Transmission rate - week 28** | Uniform [0,10] | 0.617 (95%CI: 0.572-0.667) | 2140 | 1.03 |
| **Transmission rate - week 29** | Uniform [0,10] | 0.599 (95%CI: 0.552-0.648) | 2270 | 1.01 |
| **Transmission rate - week 30** | Uniform [0,10] | 0.565 (95%CI: 0.522-0.615) | 1550 | 1.02 |
| **Transmission rate - week 31** | Uniform [0,10] | 0.529 (95%CI: 0.488-0.575) | 1150 | 1.02 |
| **Transmission rate - week 32** | Uniform [0,10] | 0.477 (95%CI: 0.438-0.521) | 1340 | 1.02 |
| **Transmission rate - week 33** | Uniform [0,10] | 0.409 (95%CI: 0.376-0.448) | 2020 | 1.05 |
| **Transmission rate - week 34** | Uniform [0,10] | 0.411 (95%CI: 0.377-0.451) | 1140 | 1.03 |
| **Transmission rate - week 35** | Uniform [0,10] | 0.424<br>(95%CI:<br>0.387-0.465) | 1470 | 1.05 |
| **Transmission rate - week 36** | Uniform [0,10] | 0.395<br>(95%CI:<br>0.359-0.436) | 1600 | 1.03 |
| **Transmission rate - week 37** | Uniform [0,10] | 0.376<br>(95%CI:<br>0.341-0.417) | 1260 | 1.03 |
| **Transmission rate - week 38** | Uniform [0,10] | 0.375<br>(95%CI:<br>0.338-0.416) | 1270 | 1.03 |
| **Transmission rate - week 39** | Uniform [0,10] | 0.342<br>(95%CI:<br>0.305-0.383) | 1570 | 1.02 |
| **Transmission rate - week 40** | Uniform [0,10] | 0.356<br>(95%CI:<br>0.317-0.401) | 1880 | 1.03 |
| **Transmission rate - week 41** | Uniform [0,10] | 0.321<br>(95%CI:<br>0.282-0.363) | 1390 | 1.02 |
| **Transmission rate - week 42** | Uniform [0,10] | 0.312<br>(95%CI:<br>0.271-0.36) | 1370 | 1.02 |
| **Transmission rate - week 43** | Uniform [0,10] | 0.283<br>(95%CI:<br>0.241-0.333) | 1500 | 1.01 |
| **Transmission rate - week 44** | Uniform [0,10] | 0.246<br>(95%CI: 0.203-0.296) | 2210 | 1.01 |
| **Transmission rate - week 45** | Uniform [0,10] | 0.248<br>(95%CI: 0.2-0.306) | 1650 | 1 |
| **Transmission rate - week 46** | Uniform [0,10] | 0.277<br>(95%CI: 0.218-0.347) | 1950 | 1.01 |
| **Transmission rate - week 47** | Uniform [0,10] | 0.231<br>(95%CI: 0.171-0.304) | 3870 | 1 |
| **Transmission rate - week 48** | Uniform [0,10] | 0.209<br>(95%CI: 0.145-0.291) | 2870 | 1 |
| **Transmission rate - week 49** | Uniform [0,10] | 0.188<br>(95%CI: 0.118-0.282) | 3320 | 1 |
| **Transmission rate - week 50** | Uniform [0,10] | 0.226<br>(95%CI: 0.135-0.348) | 1830 | 1 |
| **Transmission rate - week 51** | Uniform [0,10] | 0.244<br>(95%CI: 0.139-0.394) | 3310 | 1 |
| **Transmission rate - week 52** | Uniform [0,10] | 0.221<br>(95%CI: 0.109-0.395) | 2770 | 1 |
| **Transmission rate - week 53** | Uniform [0,10] | 0.206<br>(95%CI:<br>0.0884-0.423) | 4550 | 1 |
| **Transmission rate - week 54** | Uniform [0,10] | 0.161<br>(95%CI:<br>0.047-0.409) | 4640 | 1 |
| **Transmission rate - week 55** | Uniform [0,10] | 0.238<br>(95%CI:<br>0.0703-0.578) | 3540 | 1 |

**Figure S1:**
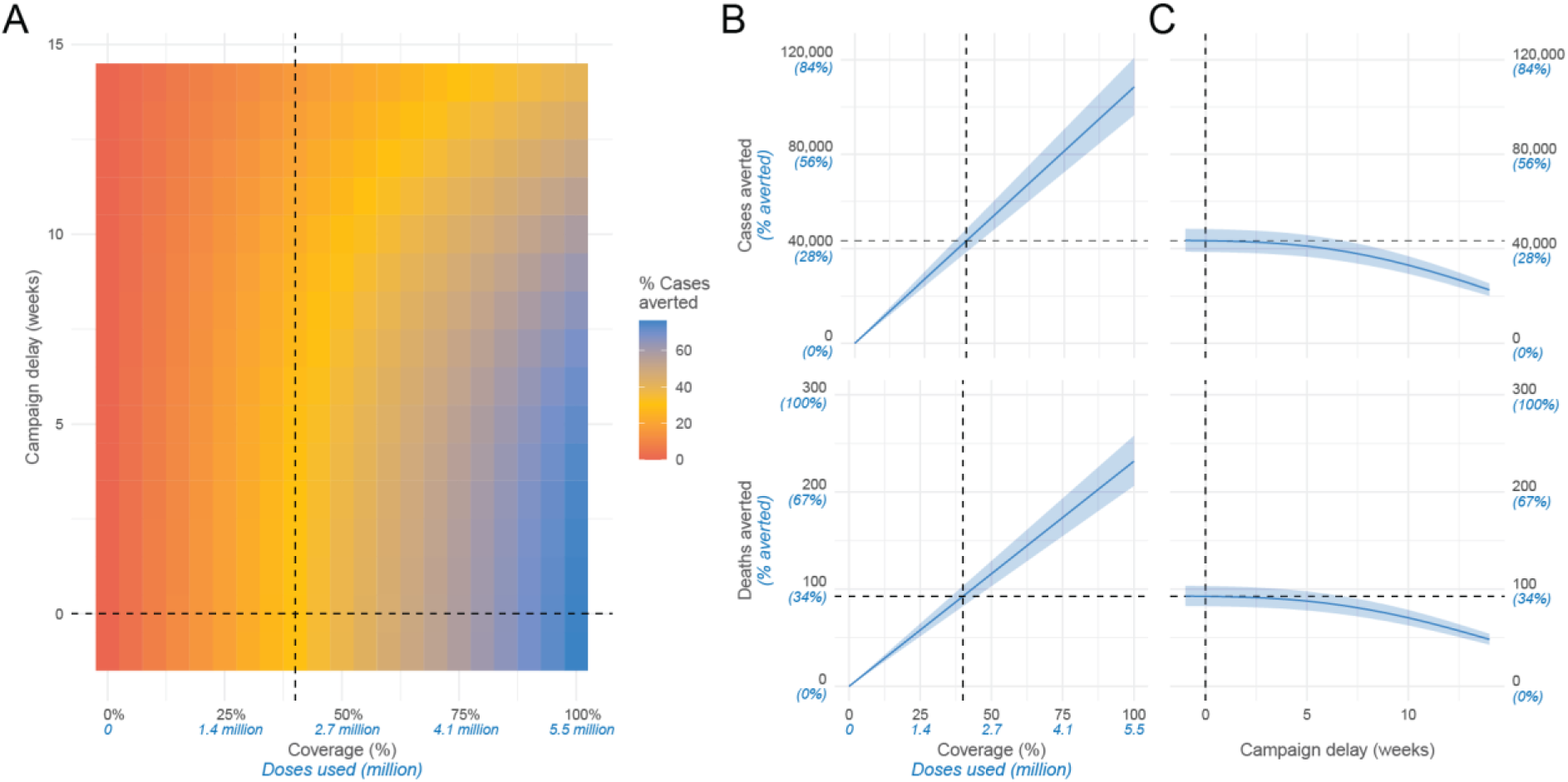
Results of vaccine model - sensitivity with a 98% vaccine efficacy. **(A)** Proportion of cases averted for different values of coverage and delay. The dashed black line shows the base case scenario of 40% coverage with no delay between outbreak start and campaign vaccination. **(B)** Infections (top row), cases (middle row) and deaths (bottom row) averted when varying coverage and delay are fixed at 0 weeks. **(C)** Infections (top row), cases (middle row) and deaths (bottom row) averted when varying the delay between the detection of the outbreak and the start of the vaccine campaign, fixing coverage at 40%.

**Figure S2:**
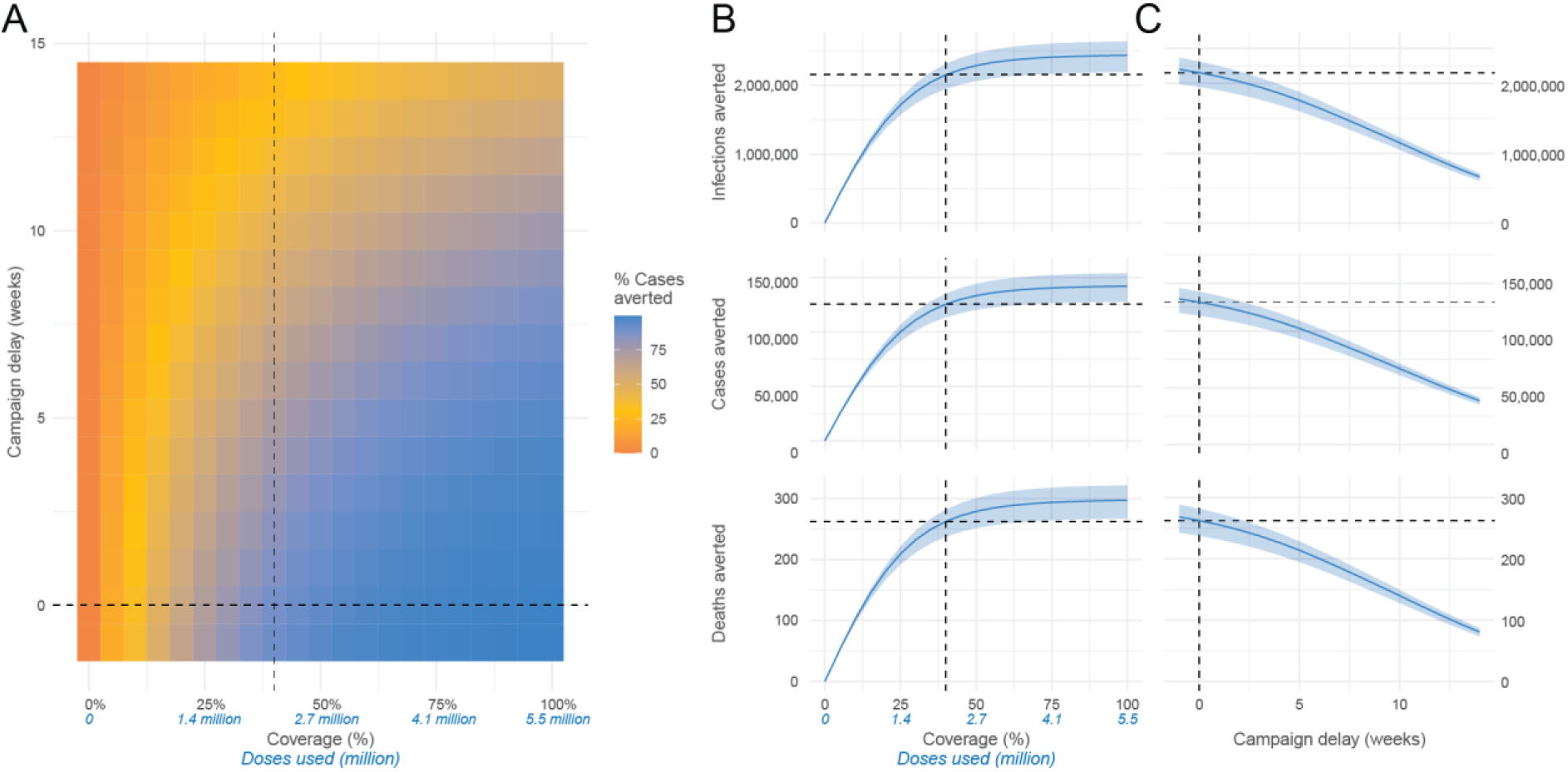
Results of vaccine model - sensitivity with infection-blocking vaccine. **(A)** Proportion of cases averted for different values of coverage and delay. The dashed black line shows the base case scenario of 40% coverage with no delay between outbreak start and campaign vaccination. **(B)** Infections (top row), cases (middle row) and deaths (bottom row) averted when varying coverage and delay are fixed at 0 weeks. **(C)** Infections (top row), cases (middle row) and deaths (bottom row) averted when varying the delay between the detection of the outbreak and the start of the vaccine campaign, fixing coverage at 40%.

**Figure S3:**
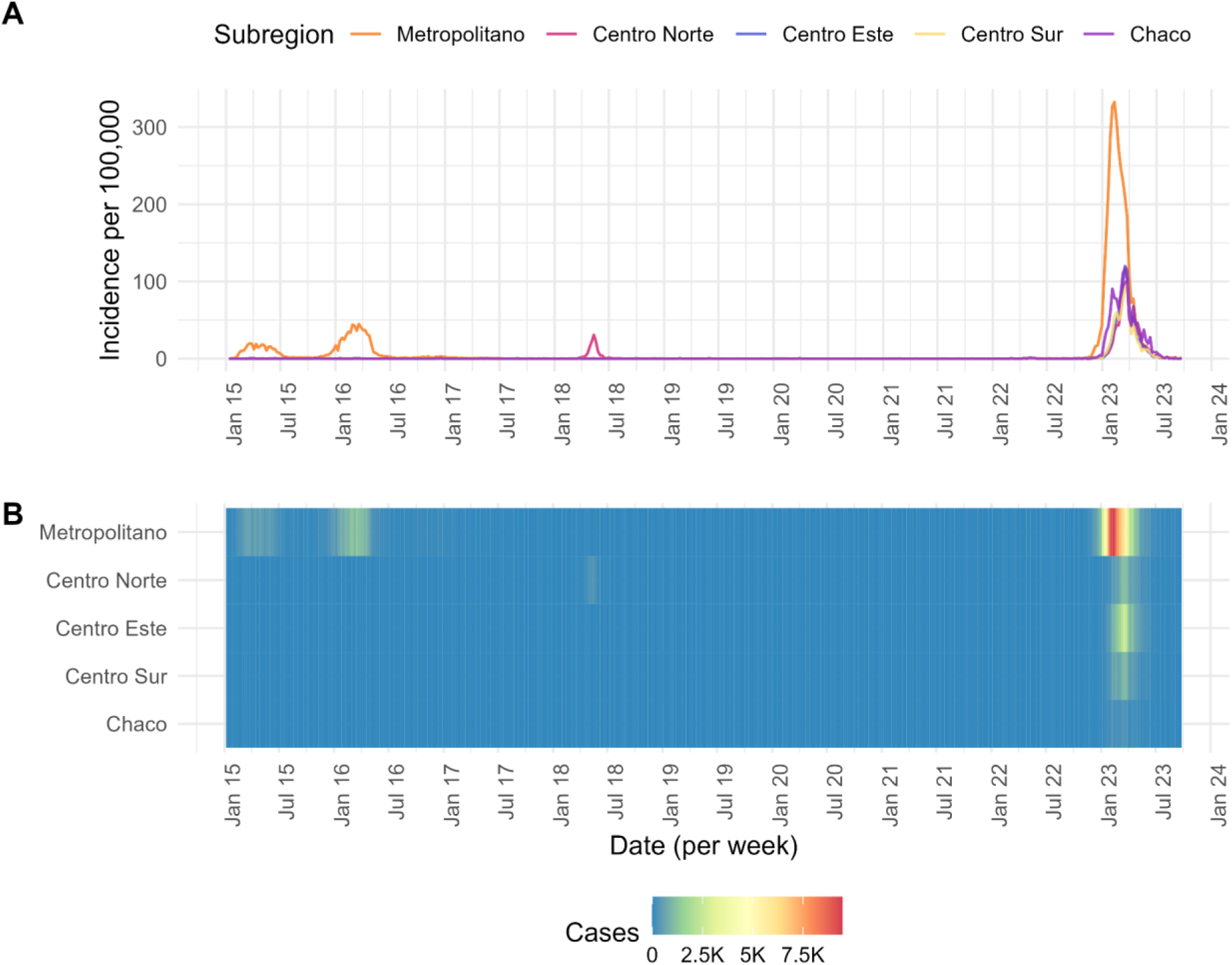
**(A)** Chikungunya cases per week in Paraguay subregions between 2015-2023. **(B)** Cases reported on the same dates.

**Figure S4.**
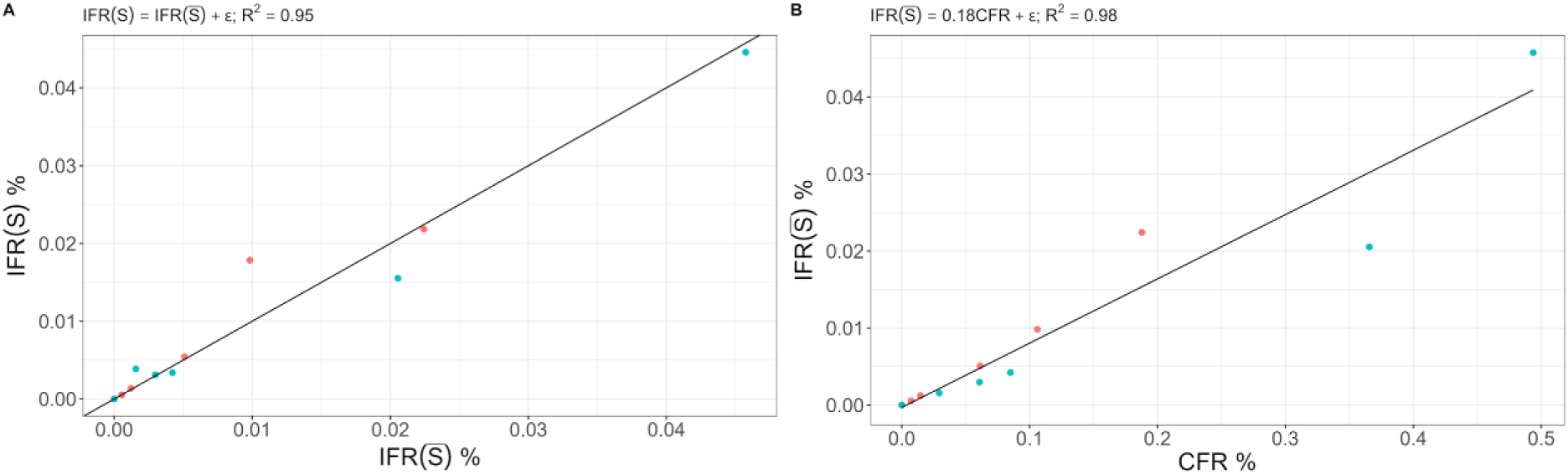
Impact of assuming equal infection risk across age and sex groups. **(A)** Infection fatality ratio (IFR) calculated using overall seroprevalence (i.e., where the same attack rate is assumed for all age/sex groups) vs IFR calculated using age and sex-specific seroprevalence (i.e., where we use age/sex specific attack rates from the seroprevalence study, where this is available). Red dots are females, blue dots are males. **(B)** Infection fatality ratio (IFR) calculated using overall seroprevalence <u>*S*</u> against Case Fatality Ratio (CFR) calculated using age and sex-specific seroprevalence. Red dots are females, blue dots are males. The lines represent the results of a linear regression.

## Acknowledgements

Funding acknowledgements section

The authors would like to acknowledge funding from Coalition for Epidemic Preparedness Innovations (CEPI) and the European Research Council (grant number 804744).

## Author contributions

Conceived study: PP-E, GRdS, HS; Wrote initial draft: PP-E, GRdS, HS; Coordination and testing of seroprevalence study: MJO, GS, LL, CF, EV, AC,PP-E, CV, AKI-V, GS, SV; Data analysis: SC, HS, PP-E, GRdS; Expert input on model inputs and data interpretation: JLDF, DS, CM, AD Data procurement: AC, EV; Revision of manuscript: All authors.

## Data and code availability

[The data from the seroprevalence study will be made available as part of this paper, with personal identifiers removed. All code and data necessary to recreate the analyses will be available through a GitHub repository https://github.com/Pastor-E-Perez-Estigarribia/ePY.]

## Ethical approval

The seroprevalence study was conducted as part of the Paraguay Ministry of Health Public Health Emergency Response and, therefore, did not require ethical approval. The seroprevalence study has not been published (either in full or in part) elsewhere. Following a standard operating process, the “Centro Nacional de Servicios de Sangre” (CENSSA), with the signed authorisation of the responsible Director, provided all the donated blood samples in this study. All blood samples were collected anonymously, with no identifiers that could link the sample to the original patient; there was no enrolment procedure, and the patient consent form was the standard used by the “Centro Nacional de Servicios de Sangre” (CENSSA). The biochemical analyses were conducted at the Central Public Health Laboratory of the Ministry of Health in Paraguay. The use of epidemiological surveillance data was authorised by the signature of the Director of the DGVS (Dirección General de Vigilancia de la Salud). The mathematical modelling work was conducted on aggregate data and, therefore, did not constitute personally identifiable information.

## References

1. Weaver, S. C. & Forrester, N. L. Chikungunya: Evolutionary history and recent epidemic spread. Antiviral Res. 120, 32–39 (2015).

2. Kang, H. et al. Chikungunya seroprevalence, force of infection, and prevalence of chronic disability after infection in endemic and epidemic settings: a systematic review, meta-analysis, and modelling study. Lancet Infect. Dis. 24, 488–503 (2024).

3. O’Driscoll, M., Salje, H., Chang, A. Y. & Watson, H. Arthralgia resolution rate following chikungunya virus infection. Int. J. Infect. Dis. 112, 1–7 (2021).

4. Salje, H. & Cortés Azuero, O. The deadly potential of chikungunya virus. Lancet Infect. Dis. 24, 442–444 (2024).

5. de Souza, W. M. et al. Spatiotemporal dynamics and recurrence of chikungunya virus in Brazil: an epidemiological study. Lancet Microbe 4, e319–e329 (2023).

6. Harris, E. FDA Approves First Chikungunya Vaccine. JAMA 330, 2241 (2023).

7. More than US$ 1.8 billion in support for African vaccine manufacturing, catching up missed children and pandemic preparedness approved as Gavi Board steps up efforts to tackle backsliding and fight health emergencies. https://www.gavi.org/news/media-room/initiatives-african-vaccine-manufacturing-approved-gavi-board (2023).

8. Desai, S. N. et al. Achievements and challenges for the use of killed oral cholera vaccines in the global stockpile era. Hum. Vaccin. Immunother. 13, 579–587 (2017).

9. Kuno, G. A Re-Examination of the History of Etiologic Confusion between Dengue and Chikungunya. PLoS Negl. Trop. Dis. 9, e0004101 (2015).

10. Yoon, I.-K., et al. High rate of subclinical chikungunya virus infection and association of neutralizing antibody with protection in a prospective cohort in the Philippines. PLoS Negl. Trop. Dis. 9, e0003764 (2015).

11. Kyungah Lim, J., et al. Seroepidemiological reconstruction of long-term chikungunya virus circulation in Burkina Faso and Gabon. J. Infect. Dis. jiac246 (2022).

12. Giovanetti, M. et al. Rapid Epidemic Expansion of Chikungunya Virus East/Central/South African Lineage, Paraguay. Emerg. Infect. Dis. 29, 1859–1863 (2023).

13. Sequera, G. ¿Por qué esta gran epidemia de Chikungunya? ¿Qué paso del Dengue? An. Fac. Cienc. Méd. (Asunción) 56, 19–24 (2023).

14. Gutiérrez, L. A. PAHO/WHO Data - Weekly Report. Pan American Health Organization / World Health Organization https://www3.paho.org/data/index.php/en/mnu-topics/chikv-en/550-chikv-weekly-en.html (2019).

15. Yoon, I.-K. et al. Pre-existing chikungunya virus neutralizing antibodies correlate with risk of symptomatic infection and subclinical seroconversion in a Philippine cohort. Int. J. Infect. Dis. 95, 167–173 (2020).

16. Kumar, M. S. et al. Seroprevalence of chikungunya virus infection in India, 2017: a cross-sectional population-based serosurvey. Lancet Microbe 2, e41–e47 (2021).

17. Salje, H. et al. Reconstruction of 60 Years of Chikungunya Epidemiology in the Philippines Demonstrates Episodic and Focal Transmission. J. Infect. Dis. 213, 604–610 (2016).

18. Schneider, M. et al. Safety and immunogenicity of a single-shot live-attenuated chikungunya vaccine: a double-blind, multicentre, randomised, placebo-controlled, phase 3 trial. Lancet 401, 2138–2147 (2023).

19. de Souza, W. M. et al. Chikungunya: a decade of burden in the Americas. Lancet Reg. Health Am. 30, 100673 (2024).

20. Salje, H. et al. How social structures, space, and behaviors shape the spread of infectious diseases using chikungunya as a case study. Proc. Natl. Acad. Sci. U. S. A. 113, 13420–13425 (2016).

21. Hozé, N. et al. Reconstructing Mayaro virus circulation in French Guiana shows frequent spillovers. Nat. Commun. 11, 2842 (2020).

22. Metcalf, C. J. E. et al. Comparing the age and sex trajectories of SARS-CoV-2 morbidity and mortality with other respiratory pathogens. R Soc Open Sci 9, 211498 (2022).

23. Torales, M. et al. Notes from the Field: Chikungunya Outbreak - Paraguay, 2022-2023. MMWR Morb. Mortal. Wkly. Rep. 72, 636–638 (2023).

24. Chen, L. H., Fritzer, A., Hochreiter, R., Dubischar, K. & Meyer, S. From bench to clinic: the development of VLA1553/IXCHIQ, a live-attenuated chikungunya vaccine. J. Travel Med. 31, taae123 (2024).

25. McMahon, R. et al. A randomized, double-blinded Phase 3 study to demonstrate lot-to-lot consistency and to confirm immunogenicity and safety of the live-attenuated chikungunya virus vaccine candidate VLA1553 in healthy adults. J. Travel Med. 31, (2024).

